# Efficacy and safety of tenofovir disoproxil fumarate (TDF) in hepatitis B virus (HBV) monoinfection: longitudinal analysis of a UK cohort

**DOI:** 10.1101/2020.12.11.20247940

**Authors:** Tingyan Wang, David A Smith, Cori Campbell, Jolynne Mokaya, Oliver Freeman, Hizni Salih, Anna L McNaughton, Sarah Cripps, Kinga A Várnai, Theresa Noble, Kerrie Woods, Jane Collier, Katie Jeffery, Jim Davies, Eleanor Barnes, Philippa C Matthews

## Abstract

**Aim:** Current clinical recommendations suggest treating chronic hepatitis B virus (HBV) infection in a minority of cases, but more data are needed to determine the benefits and risks of Tenofovir disoproxil fumarate (TDF) therapy. We aimed to assess the impact of TDF on liver disease, and the risk of nephrotoxicity.

**Method:** We studied a longitudinal UK chronic HBV (CHB) cohort attending out-patient clinics between 2005 and 2018, analysing data for 206 ethnically diverse adults (60 on TDF, 146 untreated), with median follow-up 3.3±2.8 years.

**Results:** Patients prescribed TDF were older (39 vs. 35 years, p=0.004) with a male excess (63% vs. 47%, p=0.04) compared to untreated patients. Reflecting treatment eligibility criteria, at baseline, treated patients were more likely to have elevated ALT (p<0.001), higher HBV DNA viral load (VL) (p<0.001), and higher elastography scores (p=0.002), but with no difference in renal function (p=0.6). In the TDF group, VL declined significantly between baseline and subsequent time points (all p<0.0001) with VL suppressed in 94% at three years, while in the untreated group viraemia was unchanged from baseline. In the TDF group, ALT and elastography scores normalised during treatment and by three years were equivalent to those in the untreated group. Progression of liver fibrosis did not occur in the TDF group but arose in 7.4% of untreated patients, although this difference was non-significant. There was no significant difference in renal impairment during follow-up between two groups.

**Conclusion:** TDF may have long-term benefits for a wider pool of the CHB population.

## INTRODUCTION

257 million people are living with chronic hepatitis B virus (HBV) infection according to World Health Organisation (WHO) estimates in 2015. Of these, only ∼27 million are aware of their HBV infection, among whom 4.5 million are on treatment (1). HBV results in close to one million deaths every year, mostly from cirrhosis and hepatocellular carcinoma (HCC). Curing chronic HBV infection (CHB) is a challenge due to the persistence of covalently closed circular DNA (cccDNA) and integrated HBV DNA (2) However, nucleot(s)ide analogues (NA) can be used to suppress viral replication, thus reducing the risk of inflammatory liver disease, cirrhosis and HCC (2, 3). HBV surface antigen (HBsAg) loss, sometimes termed ‘functional cure’, is regarded as the optimal endpoint of current treatment in clinical guidelines, but is infrequently achieved, as evidenced by the annual rate of HBsAg loss ranging from 0.12% to 2.7% (4-6). Therefore, NA therapy is typically continued long-term, or even life-long (2).

Tenofovir disoproxil fumarate (TDF) is affordable, well tolerated and has a high genetic barrier to resistance, and is thus the most commonly used first-line NA option for patients with CHB. However, existing guidelines (2, 3, 7) recommend that patients are stratified for therapy based on demographic, clinical and laboratory parameters, such that only a minority (e.g., ranging from 2%∼31% in different settings) (8, 9) are treatment eligible. Long-term efficacy of TDF has been confirmed by long-term follow up of treated CHB patients, showing viral suppression (10), improvements in liver histology in patients with baseline cirrhosis (11, 12), and reduced cumulative probability of HCC, hepatic decompensation, death and liver transplantation, compared to non-treatment (13). A retrospective study of >29,000 CHB adults in China (among whom a low proportion had cirrhosis at baseline) treated with entecavir (ETV) or TDF for at least six months (median follow-up 3.6 years) found that TDF treatment was associated with a lower risk of HCC than ETV treatment (14). Although these previous studies provide evidence for the benefits of TDF treatment, they mainly focus on the association between TDF and HCC or cirrhosis. In contrast, there are few data on evolution or progression of liver inflammation and fibrosis in non-cirrhotic HBV patients (11).

Benefits associated with TDF therapy have to be balanced against concerns about potential risks, of which the best recognised is renal toxicity (2). However, most evidence for TDF-mediated renal injury comes from human immunodeficiency virus (HIV) field and reported associations may be confounded by other drugs, comorbidities, or HIV associated nephropathy (15, 16). HBV guidelines produced by the European Association for the Study of the Liver (EASL) recommend that all TDF treated patients should have renal monitoring (2). One study carried out among CHB patients with high HBV DNA (>6 log_10_ IU/ml) reported that TDF treatment causes a higher incidence of acute kidney injury (AKI) compared to entecavir (ETV), but the difference was only borderline significant over three years (17), and similar risks have also been documented in other cohorts (18). In the context of baseline renal impairment and diabetes mellitus, TDF may increase the risk of worsening renal function (19, 20). However, this observation is not consistent, as other studies have not identified any significant difference in the risk of renal events between TDF and ETV groups (21-23), or report no deterioration in renal function on therapy (24) particularly if there is no renal impairment at baseline (20). In summary, although the safety of TDF on HBV patients has been explored, the long-term renal safety of TDF remains controversial (17, 18, 21-24).

The varying findings regarding the risk of renal toxicity for patients on TDF therapy may be due to differences in the characteristics of patient cohorts in terms of age, estimated glomerular filtration rate (eGFR) baseline, and prevalence of other chronic health conditions such as hypertension and diabetes (25, 26). Most previous studies have focused on longitudinal comparison of CHB treated with TDF vs. non-TDF agents, but there are limited studies comparing changes in renal function over time between treated and untreated populations (27, 28). Furthermore, previous studies have typically only compared average renal function from the cohorts at each time point, rather than examining longitudinal changes in renal function in each patient.

To examine the influence of TDF on reducing inflammatory liver disease, and to assess the renal safety of TDF in CHB patients, we aimed to analyse longitudinal changes in liver enzymes, elastography, and renal function both at a population level and within individuals, comparing patients treated with TDF therapy vs. those not on HBV treatment. Specifically, we address: a) the effects of TDF treatment on maintaining viral suppression, and reversal of liver inflammation and fibrosis compared to untreated patients; and b) the risks of renal toxicity associated with TDF treatment in CHB patients with mild/moderate liver disease at baseline.

## MATERIALS AND METHODS

### Study cohort

We conducted a longitudinal retrospective study on an adult CHB cohort in the United Kingdom between 06/2005 and 10/2018. Data were collected from Oxford University Hospitals (OUH) National Health Service (NHS) Foundation Trust, a large teaching hospital trust in the South East of the UK using a clinical informatics pipeline supported by the National Institute for Health Research Health Informatics Collaborative (NIHR HIC) (6, 29).

The baseline is defined as the start date of treatment for TDF group and the beginning of clinical follow-up for the untreated group. The inclusion criteria were: (1) patients with CHB (defined as two positive HBV surface antigen (HBsAg) tests and/or detectable HBV DNA tests at least six months apart); and (2) patients on TDF monotherapy or treatment naïve patients (i.e., individuals who were not on treatment with NAs or interferon).

The exclusion criteria were: (1) patients with hepatitis C virus (HCV), hepatitis D virus (HDV) or HIV coinfection; (2) patients with decompensated cirrhosis at baseline; (3) patients who had HCC at baseline; (4) patients who were younger than 18 years at baseline; or (5) patients who had been followed-up for less than one year from baseline.

The detailed information for selecting patients based on each criterion is illustrated in a flow diagram (**Figure S1**). We included 206 patients (60 patients with TDF monotherapy, and 146 patients without treatment). All patients had at least two measurements of alanine transaminase (ALT), eGFR, and serum creatinine. 202/206 patients had at least two measurements of HBV DNA VL.

### Laboratory markers

Quantitative HBV DNA testing was undertaken at OUH NHS Foundation Trust clinical diagnostic microbiology laboratory using the Cobas TaqMan assay (Roche Diagnostics, Branchburg, NJ), with a lower limit of detection of 9 IU/ml. Serum HBV e antigen (HBeAg) was measured using Centaur (09/2004 - 12/2014) or Abbott Architect i2000SR (Abbott Laboratories, Chicago, IL) (12/2014 - 10/2018). Quantitative HBsAg was tested using Centaur (09/2004 - 12/2014) or Abbott Architect i2000SR (Abbott Laboratories, Chicago, IL) (12/2014 - 10/2018) with lower limit of detection of 0.05 IU/ml. ALT was tested using Siemens ADVIA 2400 (02/2013 - 01/2015) or Abbott Architect c16000 or c8000 (Abbott Laboratories, Chicago, IL) (01/2015 - 10/2018) with the normal reference range from 10 to 45 IU/L. There was no distinct ALT reference range for males and females. Creatinine was measured in micromoles per litre (μmol/L) in this study with the normal reference range from 64-104 umol/L.

We defined: (a) virologic response as detectable serum HBV DNA viral load (VL) at baseline which is suppressed to <20 IU/ml during the follow-up; (b) HBeAg loss as a change from being HBeAg positive to HBeAg negative; (c) HBeAg seroconversion as the presence of antibody to HBeAg (anti-HBe); (d) HBsAg loss as negative HBsAg or undetectable HBsAg by quantitative test; (e) biochemical responses as the normalisation of ALT levels (i.e. decline of baseline ALT levels to <45 IU/L and maintaining this level during the follow-up period).

### Liver fibrosis assessment

We evaluated transient elastography (TE) (or FibroScan) as a non-invasive test measuring liver stiffness in kiloPascals (kPa) as a marker of liver inflammation and/or fibrosis (30). No universally agreed thresholds are available to map TE values to histological METAVIR stages (31), but various cut-off values have been suggested in HCV and HBV populations (32, 33). We used the following cut-off values for fibrosis stages: F0 with TE score of <7.0 kPa, F1 with TE score of ≥ 7.0 kPa to <8.0 kPa, F2 with TE score of ≥ 8.0 kPa to <10.0 kPa, F3 with TE score of ≥ 10.0 kPa to <14.0 kPa, and F4 with TE score of ≥14 kPa (33).

### Renal function assessment

eGFR is used to measure renal function, and is typically classified into five categories, G1-G5, used to monitor the development or progression of chronic kidney disease (CKD) (34), as follows:

- Normal kidney function defined as eGFR ≥90 ml/min/1.73m^2^ (G1);
- Mildly decreased kidney function if eGFR in 60-89 ml/min/1.73m^2^ (G2);
- Mild-moderate loss of kidney function if eGFR in 45-59 ml/min/1.73m^2^ (G3a);
- Moderate-severe loss of kidney function if eGFR in 30-44 ml/min/1.73m^2^ (G3b);
- Severe loss of kidney function if eGFR in 15-29 ml/min/1.73m^2^ (G4);
- Kidney failure if eGFR <15 ml/min/1.73m^2^ (G5).

Adverse events related to AKI are defined as an elevation in serum creatinine of ≥0.3 mg/dL (26.5 μmol/l in this study) or ≥ to 1.5 times from a known or presumed baseline (35, 36).

### Ethnicity data

Ethnicity was originally self-reported according to NHS standard ethnic category code list (37), and we used the top-level categories, i.e., White, Mixed, Asian, Black or other ethnic groups.

### Data extraction

As data used in this study were collected via a pipeline supported by NIHR HIC and stored in a central database (29), we used structured query language (SQL) techniques for data retrieval and extraction. We focused on longitudinal changes in biomarkers, and therefore, for each patient we collected data at baseline and at multiple subsequent follow-up time points at 3, 6, 12, 18, 24, 30, 36, 42 and 48 months. As this is routinely collected clinical data, the time intervals of follow-up were not regular with missing data a certain time points. To fully use the irregular data for longitudinal change pattern analysis, we used an imputation scheme, i.e., if the data at a certain time point *t* was missing, we used the data closest to that time point which should be within *t* ± 1.5 or 3 months (**Figure S2**).

### Ethics approval

The research database for the NIHR HIC viral hepatitis theme was approved by South Central - Oxford C Research Ethics Committee (REF Number: 15/SC/0523).

### Statistical analysis

We conducted all statistical analyses in R (version: 3.6.1). Significance tests were two-sided and p values <0.05 were deemed statistically significant. We used Fisher’s exact test for categorical variables, while for continuous variables we performed Wilcoxon rank sum test. Log transformation was used for HBV DNA VL and ALT values were transformed into fold of the upper limit of normal (ULN). To examine the efficacy of TDF on VL suppression (or ALT normalisation), we compared the VL (or ALT level) at different time points after treatment to VL (or ALT level) at baseline by using paired t-test or paired Wilcoxon test properly, which depends on the size of samples or the result of normality test by Shapiro-Wilk’s test. We applied Kaplan-Meier (K-M) method to estimate the rates of virologic response and ALT normalisation over time. We used log rank test to compare the difference in the cumulative rates of HBV DNA VL suppression or ALT normalisation between subgroups. As censoring could influence interpretation on K-M curve, we marked censored subjects on the curve by points. If the event of interest (i.e., HBV DNA VL suppression, ALT normalisation) did not occur before the end of follow-up, this patient was censored (the total time to an event for this subject could not be accurately determined) (38).

To determine changing trends in TE scores, we fit linear regression lines with 95% CI on TE scores of subgroups, with reporting Pearson’s correlation coefficients and linear regression significance. We stratified TE scores into different categories for matching METAVIR stages at each time point. To examine the progression of liver fibrosis, we analysed the changes of TE score categories for individuals from baseline to the end of follow-up.

We also performed changing trend analysis using linear regression fitting for eGFR. To quantify the longitudinal variability of eGFR within individuals, we used the standard deviation (SD) of eGFR during the follow-up period for each patient, which measures the amplitudes of eGFR values from mean eGFR level within an individual. Furthermore, to examine changes in renal function, we stratified the total decline of eGFR levels during the follow-up within individuals into three categories relative to baseline or the first available observation, i.e., decline greater than 10 ml/min/1.73m^2^, decline less then 10 ml/min/1.73m^2^ and no decline. To examine the progression of CKD, we analysed the drop of eGFR categories (G1, G2, G3a, G3b, G4, G5 as defined). To assess probable AKI events, we stratified the maximum change amplitude of serum creatinine levels during the follow-up into three different elevation levels, i.e., (a) elevation ≥ 26.5 µmol/l or ≥1.5 times, (b) elevation < 26.5 µmol/l and <1.5 times, and (c) no elevation. The maximum change amplitude here is defined as the maximal value of changes in serum creatinine levels between every two consecutive tests within a patient.

## RESULTS

### Patients treated with TDF are older with more liver fibrosis compared to the untreated group at baseline

We compared baseline characteristics for individuals treated with TDF vs. non-treated (**Table 1**). There was no significant difference in the length of follow-up between the two groups (p=0.2). Treated patients were significantly older than those untreated (median 39 vs. 35 years, respectively, p=0.004), and more were male (63% vs. 47% p=0.04). As expected, based on stratification for treatment using national guidelines (7), patients treated with TDF were more likely to have raised ALT (p<0.001), higher HBV DNA VL (p<0.001), and higher elastography scores (p=0.002) at baseline. However, there was no difference in baseline renal function, based either on eGFR, urea or creatinine.

**Table 1.** Baseline characteristics of 206 adults with chronic hepatitis B virus infection in a UK hospital cohort with longitudinal electronic data collection, comparing those treated with TDF to those untreated.

|  | TDF group<br>(n=60) | Untreated group<br>(n=146) | p-value |
| --- | --- | --- | --- |
| Follow-up duration (years) | 3.8 [2.6, 4.6] | 3.0 [1.9, 4.9] | 0.176 |
| Age (years) | 39 [33, 48] | 35 [30, 40] | **0.004** |
| Age >60 (years) (%) | 4 (6.7) | 5 (3.4) | 0.510 |
| Gender (%) |  |  |  |
| Female | 20 (33.3) | 71 (48.6) | 0.064 |
| Male | 38 (63.3) | 68 (46.6) | **0.042** |
| Unreported | 2 (3.3) | 7 (4.8) | 1 |
| Ethnicity (%) <sup>§</sup> |  |  |  |
| Asian | 33 (55.0) | 33 (22.6) | **<0.001** |
| Black | 11 (18.3) | 31 (21.2) | 0.780 |
| White | 9 (15.0) | 28 (19.2) | 0.610 |
| Other | 4 (6.7) | 10 (6.8) | 1 |
| Unreported | 3 (5.0) | 44 (30.1) | **<0.001** |
| HBeAg positive (%) | 5 (8.3) | 4 (2.7) | 0.126 |
| Diabetes (%) <sup>†</sup> |  |  |  |
| Yes | 2 (3.3) | 6 (4.1) | 1 |
| No | 25 (41.7) | 72 (49.3) | 0.398 |
| Unknown | 33 (55.0) | 68 (46.6) | 0.344 |
| ALT (x ULN) (IU/L) | 1.1 [0.7, 1.8] | 0.6 [0.4, 0.8] | **<0.001** |
| HBV DNA (log<sub>10</sub> IU/ml) | 4.8[4.1, 6.1] | 2.7 [1.9, 3.5] | **<0.001** |
| qHBsAg (log<sub>10</sub> IU/ml) | 3.0 [3.0, 3.6] | 3.0 [3.0, 3.6] | 0.425 |
| Liver stiffness (kPa) <sup>‡</sup> | 7.5 [5.0, 10.0] | 5.0 [4.0, 6.0] | **0.002** |
| <8 kPa | 11 (18.3) | 69 (47.3) | **<0.001** |
| ≥ 8 kPa and <14kPa | 9 (15.0) | 6 (4.1) | **0.015** |
| ≥14 kPa | 2 (3.3) | 0 (0.0) | **<0.001** |
| Unavailable | 38 (63.3) | 71 (48.6) | 0.077 |
| eGFR (ml/min/1.73m<sup>2</sup>)<sup>#</sup> | 90 [89, >90] | 90 [87, >90] | 0.613 |
| ≥ 90 ml/min/1.73m<sup>2</sup> | 29 (48.3) | 72 (49.3) | 1 |
| 60~89 ml/min/1.73m<sup>2</sup> | 11 (18.3) | 30 (20.5) | 0.865 |
| <60 ml/min/1.73m<sup>2</sup> | 0 (0) | 1 (0.97) | 1 |
| Unavailable | 20 (33.3) | 43 (29.5) | 0.702 |
| Serum creatinine (μmol/L) | 72 [57, 80] | 73 [57, 83] | 0.766 |
| Serum urea (mmol/L) | 5 [4, 6] | 4 [4, 6] | 0.199 |
| Albumin (g/L) | 42 [38, 44] | 41 [38, 45] | 0.861 |
| ALP (IU/L) | 114 [63, 163] | 96 [63, 150] | 0.266 |
| Bilirubin (total) (umol/L) | 10 [8, 15] | 9 [7, 12] | 0.104 |
| Platelet count (x 10<sup>9</sup>/L) | 205 [162.5, 240.0] | 219 [185.5, 252.2] | 0.078 |

Data are the median [interquartile range] or number (%) unless otherwise indicated. For categorical variables, Fisher exact test was performed for comparison on cells with small counts (<5), otherwise Chi-square test was used. For continuous variables, Wilcoxon test was used for comparison due to non-normality. *p* values <0.05 were deemed statistically significant, marked in bold. <sup>§</sup> Ethnicity was originally self-reported by patients according to NHS standard ethnic categories. <sup>†</sup> Diabetes was diagnosed using glycated haemoglobin (HbA1c), an HbA1c of 6.5% or 47.5 mmol/mol is a typical threshold for diagnosing diabetes (39). <sup>‡</sup> Liver stiffness was measured by transient elastography score in kiloPascals (kPa). <sup>#</sup> For eGFR, if a level was greater than 90 ml/min/1.73m<sup>2</sup>, it was not quantified by the hospital laboratory system. ALT, Alanine aminotransferase; eGFR, estimated Glomerular Filtration Rate; ALP, Alkaline phosphatase; TDF, Tenofovir disoproxil fumarate; qHBsAg, quantitative HBsAg level; ULN, upper limit of normal.

Interestingly, a higher TDF treatment rate was observed in Asian patients (55% vs. 22.6%, p<0.001) compared to patients from other ethnic groups (**Table 1**). With analysis stratified by ethnicity (**Table S1**), we found a significantly higher proportion of patients aged >60 years, significantly higher ALT levels (p=0.01), and HBV DNA VL (p=0.02) at baseline in Asian patients, compared to patients with other ethnicities. These findings may be suggestive of a higher HBeAg-positive rate amongst Asian patients but HBeAg status was missing in 14.3%-40.9% of cases (Table S1), limiting our ability to make meaningful comparisons.

### The majority of patients on TDF treatment suppressed VL within 12 months, while treatment-naive individuals maintained a virological set point during follow-up

In the TDF group, a significant decline was observed in a pairwise comparison of VL at baseline compared to all later time points (all p<0.0001, **Figure 1A**). In the untreated group, a significant difference was only found in the pairwise comparison of VL at selected later time points (at 18, 24, 30, 42 months) compared to baseline VL (**Figure 1A**).

**Figure 1.**
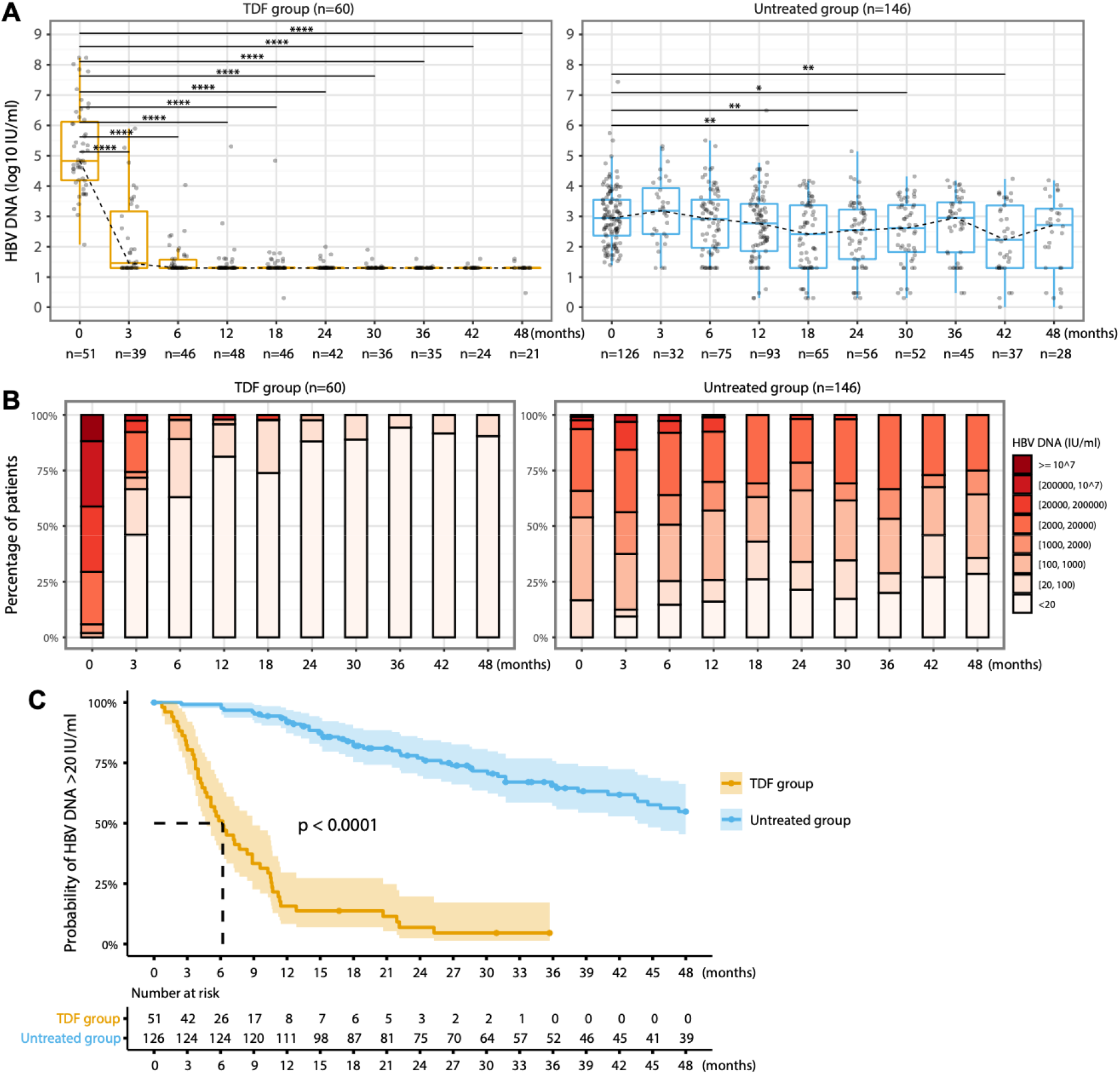
Longitudinal analysis of HBV DNA VL in chronic HBV patients with TDF treatment vs. without treatment. (A) Distribution of HBV DNA VL at each time point; (B) Stratification of HBV DNA VL at each time point; (C) Kaplan-Meier analysis to estimate the probability of patients with HBV DNA VL >20 IU/ml in each group over time. *In Kaplan-Meier analysis, real dates were used for calculating the time to HBV DNA VL suppressed <20 IU/ml, rather than the imputed time points. * p-value <0*.*05, ** p-value <0*.*01, *** p-value <0*.*001, **** p-value <0*.*0001. HBV, hepatitis B virus; Tenofovir disoproxil fumarate; VL, viral load*.

In the TDF group, 81.3% of patients suppressed VL to <20 IU/ml at one year, 88.1% at two years, and 94.1% at three years, while all had VL <2 log_10_ IU/ml at the end of follow-up (**Figure 1B**). In contrast, only ∼25% of untreated patients had VL<20 IU/ml at one to three years with the median (±IQR) is 2.8±1.5 log_10_ IU/ml, 2.6±1.6 log_10_ IU/ml, and 3.0±1.6 log_10_ IU/ml at 1 year, 2 years, and 3 years, respectively, and at the end of follow-up only 36% had VL <2 log_10_ IU/ml (**Figure 1B**).

Using a Kaplan-Meier analysis, significant differences were observed in time to virologic suppression (to <20 IU/ml) between TDF and untreated groups (p<0.0001) (**Figure 1C**). 51/60 (85%) patients in TDF group vs. 126/146 (86.3%) patients in untreated group had baseline VL data (p=0.8), and all these patients had VL>20 IU/ml at baseline. Among these patients, 94.1% (48/51) in the TDF group vs. 35.2% (44/125) in the untreated group had suppressed VL to <20 IU/ml during follow-up (p<0.001).

An increase of VL by >1 log_10_ IU/ml during follow-up was observed in 2/60 (3.3%) in the TDF group vs. 25/146 (17.1%) in the untreated group (p=0.006) (**Figure S3 and Figure S4)**. In the this untreated group, there is evidence of a set-point HBV DNA VL, where within-patient variation of VL over time accounted for 17.3% but inter-patient variation accounted for 82.7%, as previously described (40).

### Patients treated with TDF have higher baseline ALT levels that normalise with treatment

In the TDF group, ALT levels progressively normalised after treatment initiation, and significant differences were observed at six months and at later timepoints with a pairwise comparison to baseline ALT levels (all p<0.01, **Figure 2A**). In the untreated group, no differences were found for ALT at any later timepoints compared to baseline levels (**Figure 2A**). In the treated group, >70% of patients had ALT levels within the normal range at 12 months and later timepoints, while ∼85% in untreated group had normal ALT level at each time point (**Figure 2B**).

**Figure 2.**
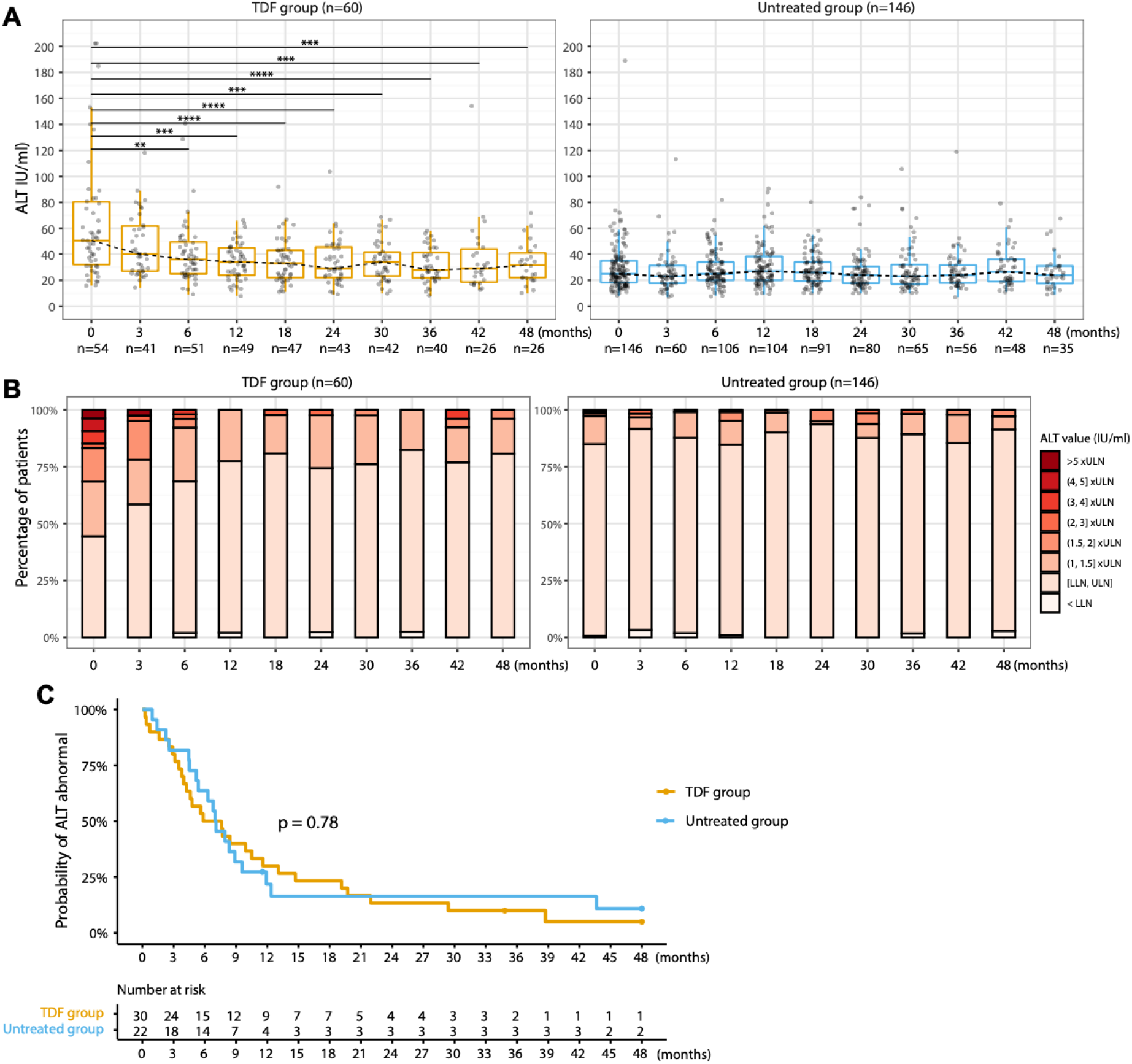
Longitudinal analysis of ALT of chronic HBV patients with TDF treatment vs. without treatment. **(A) Distribution of ALT levels at each time point; (B) Stratifications of ALT levels at each time point; (C) Cumulative percentage of patients with ALT normalisation over time**. *In Kaplan-Meier analysis, real dates were used for calculating the time to ALT normalisation, rather than the imputed time points. * p-value <0*.*05, ** p-value <0*.*01, *** p-value <0*.*001, **** p-value <0*.*0001. ALT, Alanine aminotransferase; TDF, Tenofovir disoproxil fumarate*.

Although the proportion of patients who had abnormal ALT levels at baseline was significantly higher in the TDF group (29/60, 48.3%) compared to the untreated group (22/146, 15.1%) (p<0.0001), ALT normalisation was observed in both these groups during follow-up, i.e., 89.7% (26/29) in treated patients and 86.4% (19/22) in untreated patients (p=1). By a Kaplan-Meier analysis, there was no significant difference in the time to ALT normalisation between the treated and untreated groups (p = 0.9) (**Figure 2C**).

### Transient elastography (TE) score regressed in patients treated with TDF and progressed in untreated patients

TE scores were available for 52 patients in the TDF treated group vs. 121 in the untreated group. Among these, 30/52 treated vs. 81/121 untreated patients had longitudinal measurements (≥2 TE scores). In the TDF group, the TE score was 7.5±5 (median±IQR) kPa at baseline, and declined to 6.0±1.5 kPa at four years, while in the untreated group the TE scores were 5.0±2.0 kPa during follow-up (**Figure 3A**). In the TDF group, elastography stage regressed over time, whilst a progression to F3 or F4 stages presented at 18, 30, 36, 42 months in the untreated group (**Figure 3B**). Using linear regression fitting, we observed TE scores significantly declining over time in the TDF group (R=-0.192, p=0.04), in contrast to the untreated group (R=-0.007, p=0.9) (**Figure 3C**).

**Figure 3.**
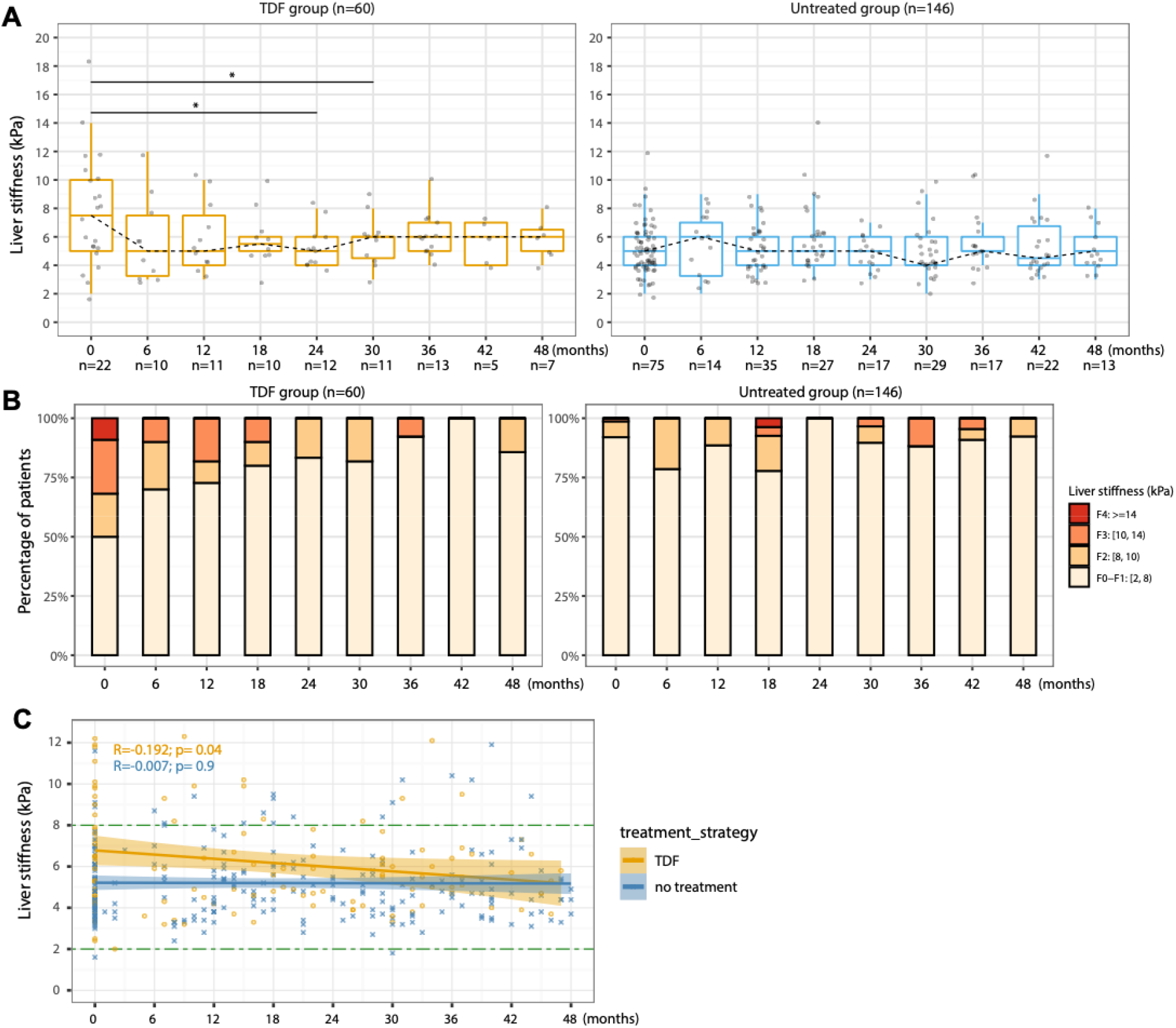
Longitudinal analysis of liver stiffness (transient elastography scores) of chronic HBV patients with TDF treatment vs. without treatment. **(A) Distribution of transient elastography scores at each time point (B) Stratifications of transient elastography scores at each time point; (C) Longitudinal changing patterns of transient elastography scores in subgroups by linear regression fitting**. *\* p-value <0*.*05, ** p-value <0*.*01, *** p-value <0*.*001, **** p-value <0*.*0001. In linear regression fitting, R represents Pearson’s correlation coefficient and p indicates linear regression significance. Tenofovir disoproxil fumarate*.

Comparing TE readings at the end of follow-up to their baseline, TE readings improved in 36.7% (11/30) of patients in TDF group (**Figure S5A**) vs. 6.2% (5/81) in the untreated group (p<0.001) (**Figure S5B**). There was no evidence of progressive liver fibrosis in the TDF group (**Figure S5A**), whereas in the untreated group, 90.1% (73/81) of patients were stage F0-F1 at baseline, of whom 6/81 (7.4%) progressed (four of these to F2 and two to F3) (**Figure S5B**). However, the difference in progression between the two groups did not reach statistical significance (0/30 vs. 6/81, p=0.2). Among untreated individuals, the baseline TE reading (median ± IQR) was 6.0±1.0 kPa in the subgroup with progressive liver fibrosis vs. 5.0±2.0 kPa in the subgroup without (p=0.09). We did not find statistically significant difference in other baseline characteristics, but due to small numbers we are underpowered to detect true associations with confidence (**Table S2**).

### No statistical difference in HBeAg and HBsAg loss rate between TDF monotherapy and non-treatment

A small number of patients (n=9) were HBeAg-positive at baseline (5 in the TDF group vs. 4 in the untreated group). HBeAg loss (seroconversion) occurred in 3/5 (60%) in the TDF group and 4/4 (100%) patients in the untreated group (one untreated patient also had HBsAg loss). HBsAg loss was recorded in 0/60 patients in the TDF group vs. 7/146 (5%) patients in the untreated group. Neither of these differences reached statistical significance (p=0.4 for HBeAg loss and p=0.2 for HBsAg loss).

### No significant difference in mild/moderate renal impairment over time between two groups and similar risks of CKD progression over time

At baseline, 11/40 treated vs. 31/103 untreated patients had eGFR <90.0 ml/min/1.73m^2^ (27.5% vs. 30.1%, p = 0.8), and 0/40 treated vs. 1/103 untreated patients had eGFR <60 ml/min/1.73m^2^ (0 vs. 1%, p = 1) (**Table 1**). During follow-up, mild renal impairment (nadir eGFR 60-89 mL/min/1.73m^2^) was present in 60% (36/60) in TDF group vs. 49% (71/146) in the untreated group (p=0.2), and moderate renal impairment (nadir eGFR 30-59 mL/min/1.73m^2^) in 5% (3/60) in TDF group vs. 3% (4/146) in untreated group (p=0.4) (**Figure 4A, B)**.

**Figure 4.**
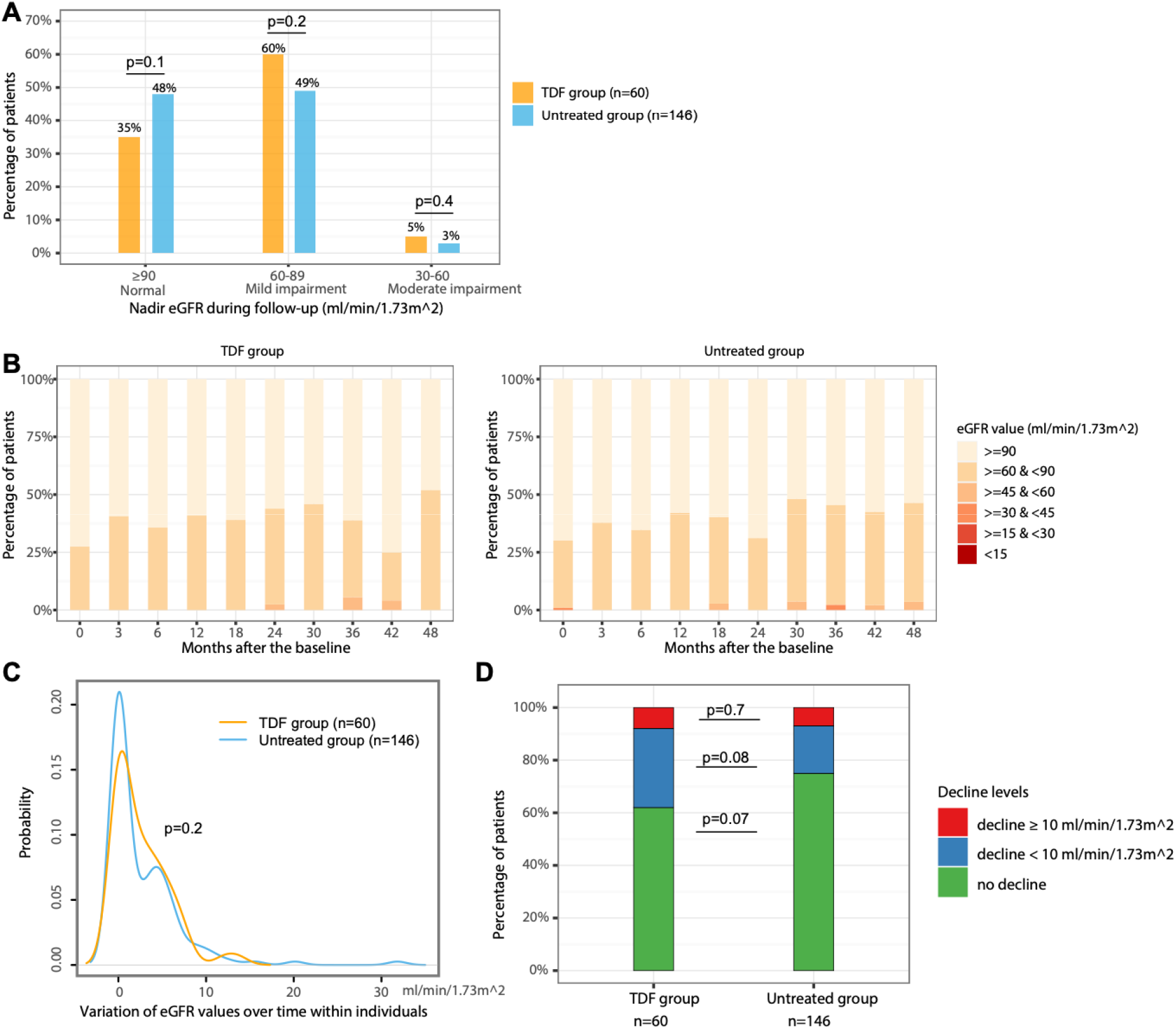
Longitudinal analysis of eGFR of chronic HBV patients with TDF treatment vs. without treatment. **(A) Comparison of the proportion of patients with different categories of nadir eGFR during follow-up between groups; (B) Distribution of CKD stages at each time point; (C) Variation of eGFR values within individuals over time; (D) Stratifications of eGFR decline levels within individuals over time**. *In panel A, nadir* eGFR *is defined as the lowest levels of eGFR of a patient during follow-up. In panel C, variation in the x-axis is calculated as the standard deviation of eGFR values of each individual during the follow-up period, y-axis is the probability of patients having a corresponding variation. eGFR, estimated Glomerular Filtration Rate; CKD, chronic kidney disease; Tenofovir disoproxil fumarate*.

Variation in eGFR within individuals over time was not significantly different between the groups (p = 0.2) (**Figure 4C**). We observed a decrease of eGFR of ≥10 mL/min/1.73m^2^ in 8% (5/60) in TDF group and 7% (10/146) of patients in untreated group (p = 0.7, red sections in **Figure 4D**), a decrease of eGFR of <10 mL/min/1.73m^2^ in 30% (18/60) in TDF group vs. 18% (26/146) in untreated group (p = 0.08, blue sections in **Figure 4D**). There was no significant difference in the proportion of patients with progression of CKD stages between the TDF group and untreated group (28.3% (17/60) vs. 17.8% (26/146), p = 0.13) (**Figure S6A, S6B)**.

### Risks of AKI events are not associated with TDF treatment

There was no significant difference in variation of serum creatinine within individuals over time between the groups (p = 0.8) (**Figure 5A**). AKI events arose in 2/60 (3.3%) of TDF-treated patients and 5/146 (3.4%) untreated patients (p=1, **Figure 5B**). We also observed a mild elevation of serum creatinine of <26.5 and <1.5 times in 95% (57/60) in TDF group vs. 81% (118/146) in untreated group (p= 0.009, **Figure 5B**), however, the median [IQR] of creatinine in both groups was within the normal reference range, i.e., 81 [69, 93] umol/L in TDF group vs. 79 [66, 89] umol/L in untreated group (p=0.2), with a small proportion who had creatinine elevated to a level >ULN (5/57 (8.8%) in TDF group vs. 5/118 (4.2%) in untreated group, p=0.3), all of these ten patients had eGFR <90.

**Figure 5.**
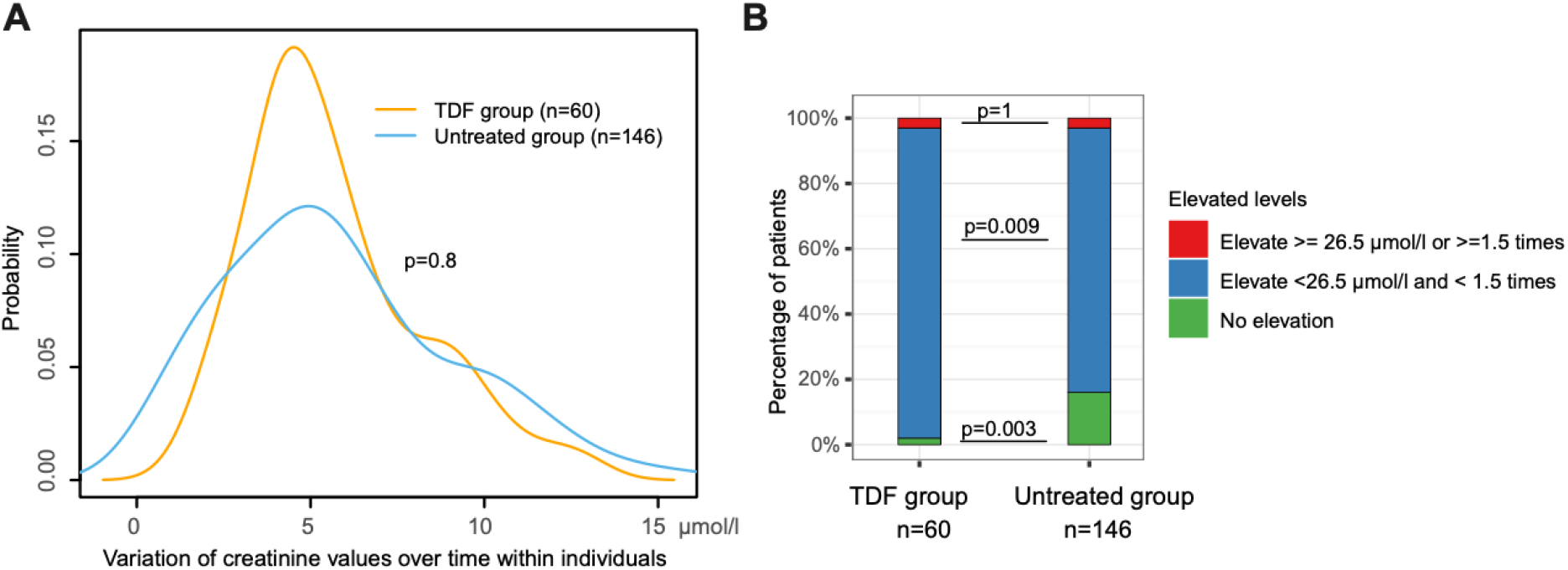
Changes of serum creatinine within individuals over time in chronic HBV patients with TDF treatment vs. without treatment. **(A) Variation of serum creatinine levels within individuals over time; (B) Stratifications of serum creatinine elevation levels within individuals over time**. *In panel A, variation in the x-axis is calculated as the standard deviation of creatinine values of an patients during the follow-up period, y-axis is the probability of patients having a corresponding variation. In panel B, red sections represent the percentage of patients had AKI events, which is defined as an elevation of serum creatinine of* ≥ *26*.*5 µmol/l or* ≥*1*.*5 times compared to previous time point. AKI, acute kidney injury. TDF, Tenofovir disoproxil fumarate*.

## DISCUSSION

### Principal findings

In this longitudinal cohort study, using an electronic pipeline developed by NIHR HIC, we show that CHB patients treated with TDF therapy suppress HBV viraemia, and this is associated with an improvement in liver stiffness when assessed up to ∼5 years. Liver disease progressed in 7.4% of patients in the untreated group over the study period and in none of the TDF treated patients. Although this difference did not reach statistical significance, this observation demonstrates evolving liver disease among a subgroup of untreated patients, highlighting that relaxation of treatment criteria could be of benefit preventing liver disease in a wider pool of the CHB population. This finding warrants exploration in larger cohorts followed up over longer time periods. Furthermore, we found no excess risk of AKI in the treated group, suggesting that extending treatment would be a low-risk strategy.

### Values added into the existing literature and clinical implications

There is currently minimal evidence assessing the development or regression of liver disease and renal function in CHB patients, receiving TDF therapy in comparison to an untreated group – especially in those without cirrhosis at baseline (11). A significantly higher treatment rate in Asian patients who had significantly higher ALT levels and HBV VL at baseline compared to other ethnicities is consistent with findings from previous studies that genotype C (common in Asia) is more likely to be HBeAg positive for longer, with higher HBV VL (41-43), and therefore Asian patients are more likely to meet current treatment criteria.

Our results provide important implications for clinical practice. Firstly, since TDF therapy is associated with disease regression in those without cirrhosis, treatment should be made more widely available to these patients. Secondly, concerns about renal toxicity should not limit access to treatment; we found that CHB without treatment also had a risk of CKD progression (consistent with a preceding cohort study (27)), which is not significantly different from the risk in treated patients. Previous studies found that risk factors associated with renal function decline in CHB patients include old age, hypertension, diabetes, baseline eGFR, and use of diuretics (44-47). Therefore, concerns about nephrotoxicity may be more pertinent in these subgroups, and risks should be assessed on a case-by-case basis.

A meta-analysis confirms that TDF and tenofovir alafenamide (TAF) are the most effective agents for virologic suppression for both HBeAg-positive and HBeAg-negative CHB patients (48), and promising short-term outcomes of TAF have been reported in several studies indicating reduced nephrotoxicity (49-51). However, a recent study of cost-effectiveness of TAF for treatment of CHB in Canada shows that TAF is not cost-effective at its current cost with a price of more than four times that of TDF, without any advantage in efficacy (52). Hence, TDF is currently considered the treatment of choice. For special groups with baseline renal dysfunction or comorbidities, however, TAF may be a safer alternative to TDF (19, 20), but long-term efficacy and safety outcomes of TAF need to be confirmed (53, 54).

### Caveats and limitations

The power of our analysis is constrained by sample size, especially for identifying associations of uncommon outcomes (e.g. loss of HBsAg or HBeAg, and progression of fibrosis in untreated patients). Although our population is ethnically diverse, the number of individuals in each group is small, and the data provides a snapshot of disease in one UK centre. We only considered adult patients and no data for special groups including pregnant women, or those with chronic comorbidities such as hypertension, diabetes, or co-infection with other blood-borne viruses. Therefore, we may have underestimated treatment impact or risk at a population level. Similar studies are needed in settings where other risk factors are more prevalent, e.g., populations where HIV is co-endemic. A study from South Africa demonstrates that adults with HIV/HBV coinfection on antiretroviral therapy have less chronic liver disease than those with HBV monoinfection, suggesting an advantage conferred by treatment in the coinfected group (55). We only compared patients on TDF treatment to untreated; due to infrequent use of ETV, we have not considered the small subgroup of our population treated with this agent. Based on the available data, we were not able to make a formal assessment of metabolic bone disease in patients on TDF therapy, and nor could we consider other reported side-effects such as gastrointestinal disturbance.

### Benefit of collecting research data in scale from EHRs

It is not possible to conduct randomized controlled trials (RCT) to study long-term outcomes of existing therapies in CHB, as it is not ethically sound not to treat those with established risk factors for pathology associated with HBV. However, there is potential to undertake studies to assess the risks and benefits of providing treatment to a wider proportion of the whole CHB population, involving patients who do not meet current criteria for eligibility. We here capitalise on the benefit of large datasets available through electronic health records (EHRs), which are routinely collected in clinical care and can be reused for research, presenting an alternative approach to large-cohort observational studies. Although data noise can be a concern in this kind of routinely collected clinical data, (e.g. irregular follow-up, missing data, changes in assay platforms and inconsistent reporting), findings will become more convincing as larger study cohorts are assimilated. Increasingly, it is possible to automate the collection of routine health care records at scale, making big data sets ready for research. As an exemplar case, the data used in this study were collected via the NIHR HIC viral hepatitis theme, and the data collection process is being rolled out to include multiple centres across the UK (29). This will also provide an opportunity to conduct validations in the real world between different centres. Consequently, it will provide more robust and reliable results since heterogeneous intrinsic characteristics exist in patients from different sources.

## Conclusion

In summary, we show clear benefits of TDF therapy in an ethnically diverse CHB population, even among patients with only mild/moderate liver disease at baseline, and suggest that these benefits may be relevant to a wider pool of the untreated CHB population without imposing clinically significant nephrotoxicity in the time frames observed here. Studies of larger populations, and over longer periods of follow-up, are urgently needed to provide the evidence to underpin expanded treatment guidelines.

## Data Availability

No additional data are available.

## Acknowledgements

This work has been supported by the National Institute for Health Research (NIHR) Biomedical Research Centre (BRC) at Oxford and funded by the NIHR Health Informatics Collaborative (HIC). C.C reports funding from GlaxoSmithKline. E.B is supported by the Oxford NIHR Biomedical Research Centre and is an NIHR Senior Investigator. P.C.M is supported by a Wellcome intermediate fellowship (grant ref 110110/Z/15/Z). The views expressed in this article are those of the author and not necessarily those of the NHS, the NIHR, or the Department of Health. The authors would like to thank all the research nurses and research admin staff at the contributing site for their help in data collection and Gail Roadknight for her work supporting the NIHR HIC.

