## Supplementary Tables and Figures for "Efficacy and safety of tenofovir disoproxil fumarate (TDF) in hepatitis B virus (HBV) monoinfection: longitudinal analysis of a UK cohort"

**SUPPLEMENTARY MATERIALS**

**Supplementary Tables**

**Supplementary Table S1**. Baseline characteristics of 206 adults with chronic hepatitis B virus infection in a UK hospital cohort stratified by patient’s ethnicity.

|  | **Asian** | **Black** | **White** | **Other** | **Unreported** | **p-value** |
| --- | --- | --- | --- | --- | --- | --- |
|  | **n=66** | **n=42** | **n=37** | **n=14** | **n=47** |  |
| Treatment status |  |  |  |  |  |  |
| TDF | 33 (50) | 11 (26.2) | 9 (24.3) | 4 (28.6) | 3 (6.4) | **<0.001** |
| Untreated | 33 (50) | 31 (73.8) | 28 (75.7) | 10 (71.4) | 44 (93.6) | **<0.001** |
| Follow-up duration (years) | 3.58 [2.50, 4.98] | 3.71 [2.46, 5.19] | 4.42 [2.83, 5.50] | 2.58 [1.65, 3.79] | 1.92 [1.50, 3.29] | **<0.001** |
| Age (years) | 39 [32, 50] | 37 [32, 41] | 35 [29, 40] | 37 [27, 44] | 33 [30, 38] | 0.06 |
| Age >60 (years) (%) | 8 (12.1) | 0 (0.0) | 0 (0.0) | 0 (0.0) | 1 (2.1) | **0.012** |
| Gender (%) |  |  |  |  |  |  |
| Female | 35 (53.0) | 15 (35.7) | 12 (32.4) | 5 (35.7) | 24 (51.1) | 0.153 |
| Male | 30 (45.5) | 25 (59.5) | 23 (62.2) | 8 (57.1) | 20 (42.6) | 0.247 |
| Unreported | 1 (1.5) | 2 (4.8) | 2 (5.4) | 1 (7.1) | 3 (6.4) | 0.536 |
| HBeAg positive (%) |  |  |  |  |  |  |
| Positive | 5 (7.6) | 1 (2.4) | 3 (8.1) | 0 (0.0) | 0 (0.0) | 0.195 |
| Negative | 34 (51.5) | 31 (73.8) | 22 (59.5) | 12 (85.7) | 39 (83.0) | **0.002** |
| Unknown | 27 (40.9) | 10 (23.8) | 12 (32.4) | 2 (14.3) | 8 (17.0) | **0.042** |
| ALT (IU/L) | 36 [24, 49] | 29 [22, 56] | 33 [21, 56] | 22 [19, 31] | 25 [19, 33] | **0.014** |
| ALT >ULN (%) ^†^ | 19 (30.2) | 11 (27.5) | 12 (32.4) | 2 (15.4) | 8 (17.0) | 0.387 |
| HBV DNA (log_10_ IU/ml) | 3.86 [2.56, 5.24] | 2.95 [1.93, 3.99] | 3.10 [2.57, 4.04] | 2.88 [2.60, 3.75] | 2.75 [2.02, 3.54] | **0.021** |

*Data are the median [interquartile range] or number (%) unless otherwise indicated. For categorical variables, Fisher exact test was performed for comparison on cells with small counts (<5), otherwise Chi-square test was used. For continuous variables, Wilcoxon test was used for comparison due to non-normality.* *p values <0.05 were deemed statistically significant, marked in bold. ^†^ 3, 2, and 1 patient(s) with Asian, black, and other ethnicity respectively had missing data on baseline ALT. ALT, Alanine aminotransferase; ULN, upper limit of normal; TDF, Tenofovir disoproxil fumarate.*

**Supplementary Table S2. Baseline characteristics of untreated patients with/without progression of liver fibrosis during follow-up.**

|  | **Untreated patients with progression of liver fibrosis**  **(n=6)** | **Untreated patients with progression of liver fibrosis**  **(n= 75)** | **p-value** |
| --- | --- | --- | --- |
| Follow-up duration (years) | 3.7 [2.1, 4.9] | 3.5 [2.17, 4.75] | 0.986 |
| Age (years) | 45 [29, 47] | 35 [30, 42] | 0.539 |
| Age >60 (years) (%) | 1 (16.7) | 4 (5.3) |  |
| Gender (%) |  |  | 1 |
| Female | 3 (50.0) | 32 (42.7) |  |
| Male | 3 (50.0) | 43 (57.3) |  |
| Ethnicity (%) ^§^ |  |  |  |
| Asian | 3 (50.0) | 13 (17.3) | 0.328 |
| Black | 1 (16.7) | 16 (21.3) |  |
| White | 0 (0.0) | 19 (25.3) |  |
| Other | 0 (0.0) | 6 (8.0) |  |
| Unreported | 2 (33.3) | 21 (28.0) |  |
| HBeAg positive (%) | 0 (0.0) | 4 (5.3) | 1 |
| Diabetes (%) ^†^ |  |  |  |
| Yes | 1 (16.7) | 2 (2.7) | 0.209 |
| No | 3 (50.0) | 39 (52.0) | 1 |
| Unknown | 2 (33.3) | 34 (45.3) | 0.688 |
| ALT (x ULN) (IU/L) | 0.7 [0.5, 0.9] | 0.6 [0.4, 0.8] | 0.482 |
| HBV DNA (log_10_ IU/ml) | 3.5 [2.8, 3.6] | 2.7 [2.0, 3.6] | 0.330 |
| qHBsAg (log_10_ IU/ml) | 3.0 [2.9, 3.5] | 3.0 [3.0, 3.6] | 0.475 |
| eGFR (ml/min/1.73m^2^) ^‡^ | 90 [87, >90] | 90 [90, >90] | 0.951 |
| Serum creatinine (μmol/L) | 67 [62, 71] | 75 [59, 81] | 0.433 |
| Serum urea (mmol/L) | 5[3, 6] | 5 [4, 6] | 0.820 |
| Liver stiffness (kPa) ^#^ | 6 [6, 7] | 5 [4, 6] | 0.089 |
| <8 kPa | 5 (83.3) | 49 (65.3) | 0.658 |
| ≥ 8 kPa and <14kPa | 0 (0.0) | 4 (5.3) | 1 |
| Unavailable | 1 (16.7) | 22 (29.3) | 0.669 |
| Albumin (g/L) | 43 [41, 44] | 42 [39, 45] | 0.864 |
| ALP (IU/L) | 108 [90, 146] | 93 [65, 150] | 0.607 |
| Bilirubin (total) (umol/L) | 8 [6, 11] | 10 [7, 13] | 0.247 |
| Platelet count (x 10^9^/L) | 253 [224, 283] | 218[181, 245] | 0.189 |

*Data are the median [interquartile range] or number (%) unless otherwise indicated. For categorical variables, Fisher exact test was performed for comparison on cells with small counts (<5), otherwise Chi-square test was used. For continuous variables, Wilcoxon test was used for comparison due to non-normality.* *p values <0.05 were marked in bold. ^§^ Ethnicity was originally self-reported by patients according to NHS standard ethnic categories. ^†^ Diabetes were diagnosed using glycated haemoglobin (HbA1c), an HbA1c of 6.5% or 47.5 mmol/mol is recommended as the cut point for diagnosing diabetes. ^‡^ For eGFR data, if a level was greater than 90 ml/min/1.73m^2^, it was originally reported as >90 by the hospital laboratory system.* ^#^ *Liver stiffness was measured by transient elastography score in kiloPascals (kPa).* *ALT, Alanine aminotransferase; eGFR, estimated Glomerular Filtration Rate; ALP, Alkaline phosphatase; qHBsAg, quantitative HBsAg level; ULN, upper limit of normal.*

**Supplementary Figures**

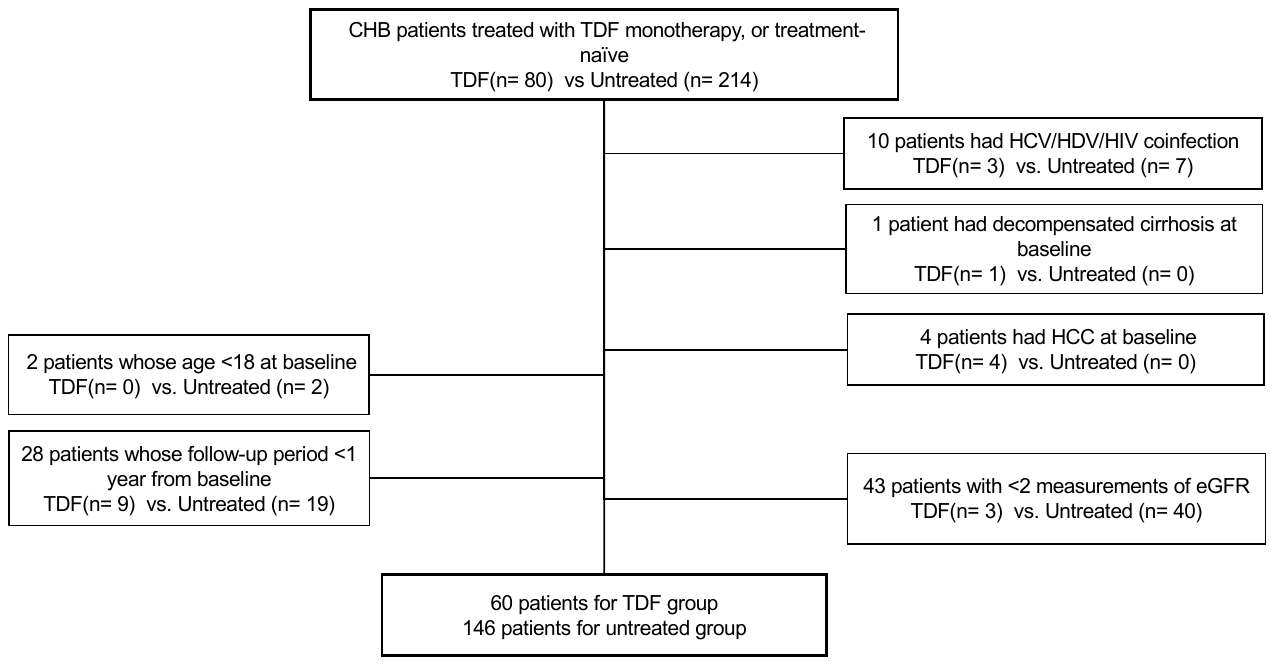

**Supplementary Figure S1. Flow chart of patients included/excluded for analysis**

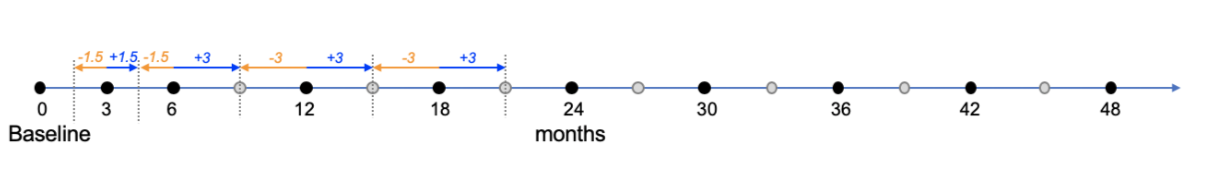

**Supplementary Figure S2. Data extraction for target time points**

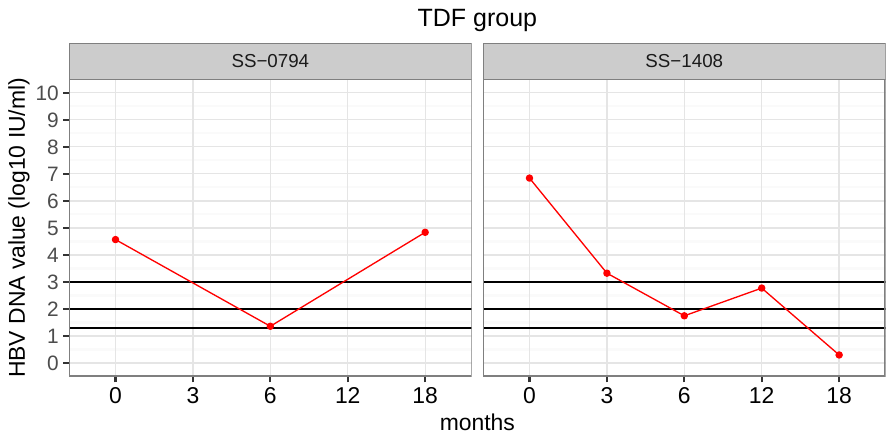

**Supplementary Figure S3. Trajectory of HBV DNA viral load for patients in TDF group with an increase of VL by 1 log10 IU/ml during follow-up.** *Each panel displays the viral load trajectory of a patient.*

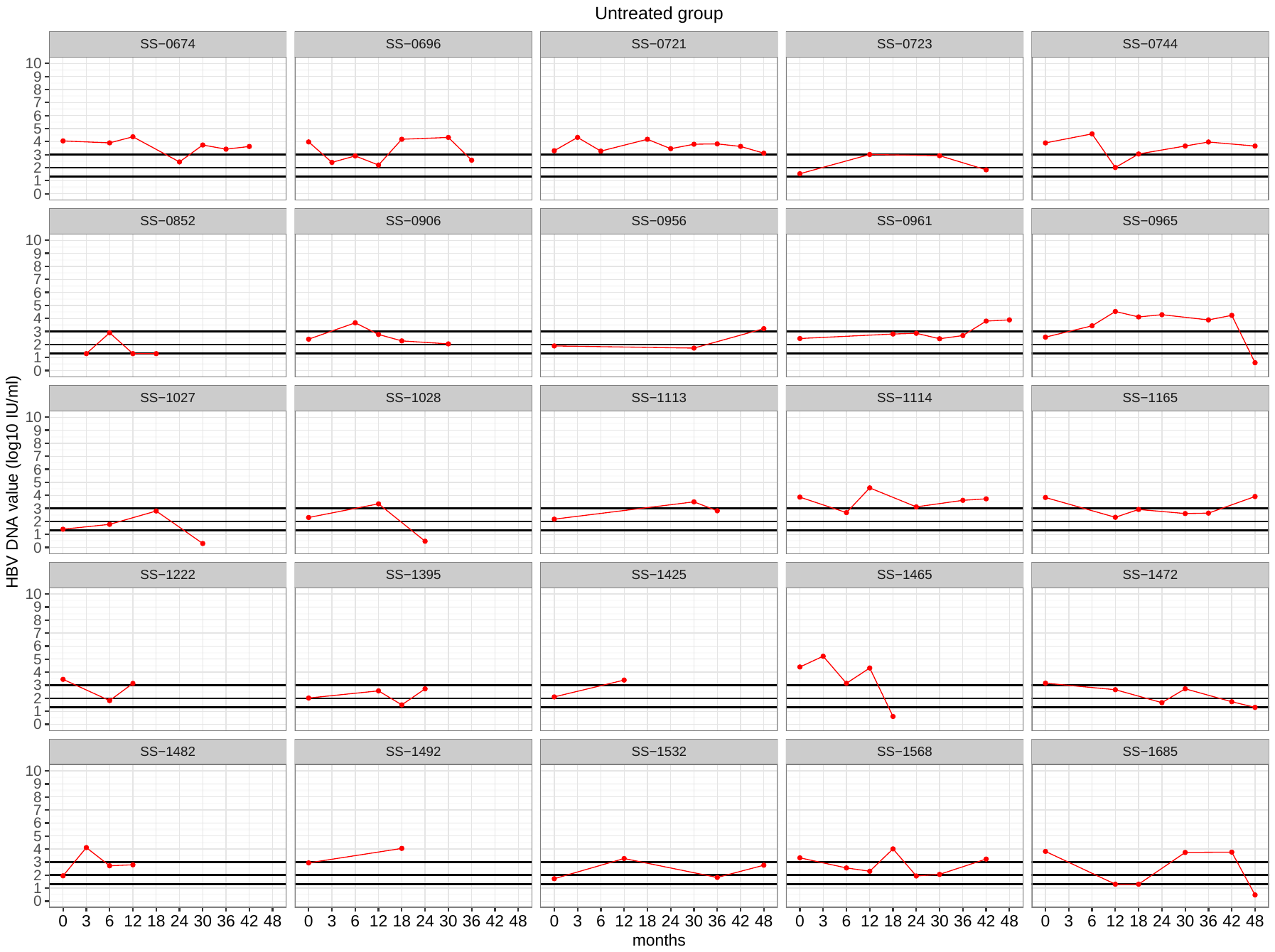

**Supplementary Figure S4. Trajectory of HBV DNA viral load for patients in untreated group with an increase of VL by 1 log10 IU/ml during follow-up.** *Each panel displays the viral load trajectory of a patient.*

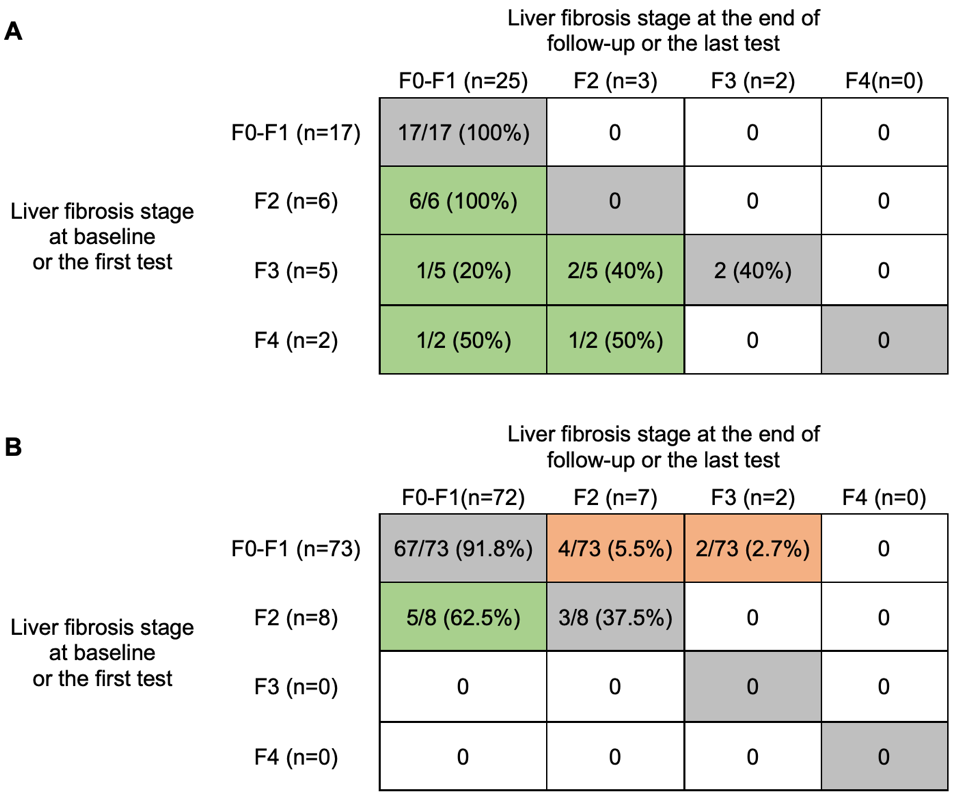

**Supplementary Figure S5.** **Changes of liver fibrosis stages from baseline to the end of follow-up for chronic HBV patients with at least two measurements of liver stiffness: (A) TDF group (n=30) (B) untreated group (n=81).** *Liver fibrosis stages were based on TE scores. F0: <7.0 kPa, F1: ≥ 7.0 kPa and <8.0 kPa, F2: ≥ 8.0 kPa and <10.0 kPa, F3: ≥ 10.0 kPa and <14.0 kPa, F4: ≥14 kPa. In TDF group, 30 patients have ≥2 measurements of TE scores, while the other 8 patients had no data of TE scores and 22 patients had only single TE score available. In untreated group, 81 patients have ≥2 measurements of TE scores, while the other 25 patients have no data of TE scores and 40 patients had only single TE score available. Cells with green colour indicate an improvement of liver stiffness, while cells with orange colour indicate a progression of liver fibrosis. Tenofovir disoproxil fumarate; TE, transient elastography.*

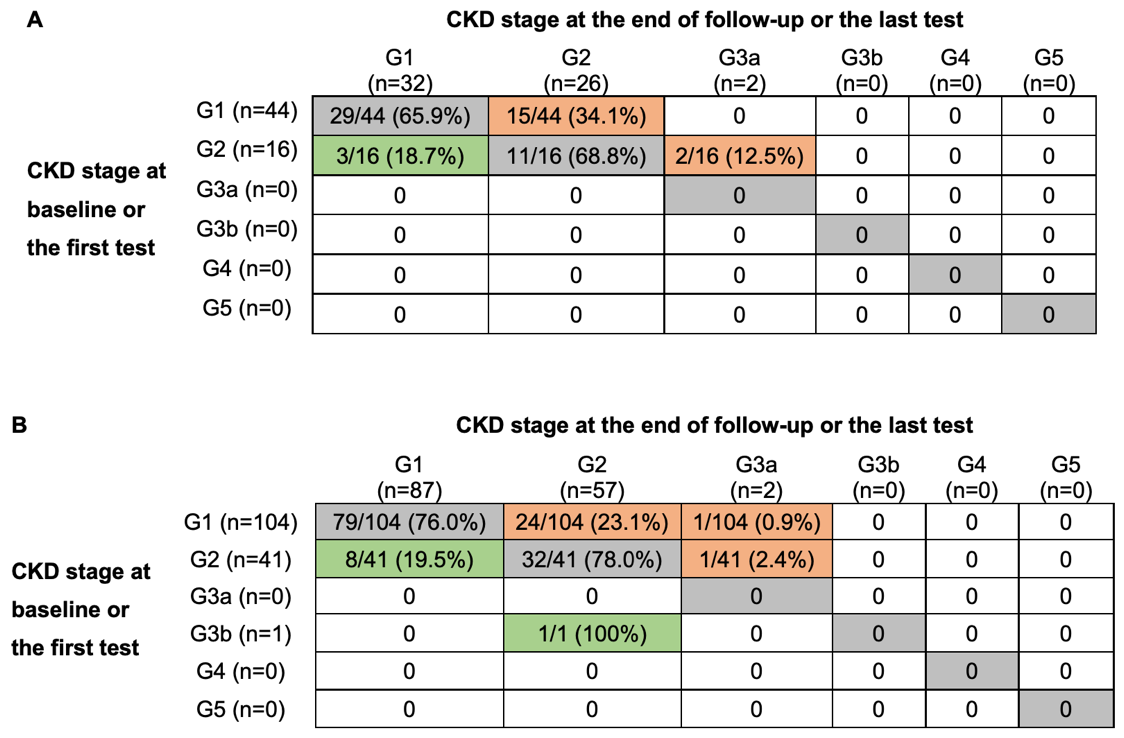

**Supplementary Figure S6. Changes of CKD stages from baseline to the end of follow-up in (A) TDF group vs. (B) untreated group.** *CKD stages were classified based on eGFR, defined as G1 (eGFR ≥90 ml/min/1.73m^2^, normal kidney function);G2 (60-89 ml/min/1.73m^2^, mildly decreased kidney function); G3a (45-59 ml/min/1.73m^2^, mild-moderate loss of kidney function); G3b (30-44 ml/min/1.73m^2^, moderate-severe loss of kidney function); G4 (15-29 ml/min/1.73m^2^, severe loss of kidney function); and G5 (eGFR <15 ml/min/1.73m^2^, kidney failure).* *Cells with green colour indicate a regression of CKD stage, while cells with orange colour indicate a progression of CKD stage. CKD, chronic kidney disease; Tenofovir disoproxil fumarate.*
